# Progenitor Cells, Regenerative Capacity, and Cognition

**DOI:** 10.1101/2025.08.26.25334500

**Authors:** Taha Ahmed, Matthew Gold, Ambar Kulshreshtha, Edward Woods, Alireza Rahbar, Mohammad Hossain, Yi-An Ko, Jose Medina-Inojosa, Kristen A. Harris, Jingwen Huang, Nodari Maisuradze, Muhammad Owais, Shaimaa M. Sakr, Tiffany Thomas, Felicia Goldstein, James Lah, Edmund Waller, Vince Calhoun, Arshed Quyyumi, Ihab Hajjar

## Abstract

**Background:** Impaired endogenous vascular regenerative capacity, reflected by reduced levels of circulating progenitor cells (CPC), has been linked to age-related diseases, especially adverse cardiovascular outcomes. We have previously reported that CPC are associated with cognitive aging, but their impact on cognitive impairment and related brain phenotypes is unclear. Here we report the impact of CPC on cognitive and neuroimaging markers of cognitive impairment.

**Methods:** We analyzed data from 283 community-dwelling participants (59% female, 39% Black) enrolled in the Brain Stress, Hemodynamics and Risk Prediction (B-SHARP) program. Participants underwent (a) cognitive assessments (including Montreal Cognitive Assessment [MoCA]); (b) brain magnetic resonance imaging (MRI) to derive white matter hyperintensity (WMH) volumes, whole-brain cortical thickness and hippocampal volumes; and (c) flow cytometry for enumerating CPCs as CD45^med^ mononuclear cells expressing CD34 with co-expression with either CD133, chemokine CXC motif receptor 4 (CXCR4), or vascular endothelial growth factor receptor-2 (VEGF2R). Linear regression models were adjusted for demographic and vascular risk factors.

**Results:** In fully adjusted models, lower levels of CD34⁺/CD133⁺ CPCs were associated with worse global cognition (MoCA: β = 0.59, p = 0.01), reduced cortical thickness (β = 0.01, p = 0.01), and greater WMH burden (β = -0.15, p = 0.01). Lower levels of CD34⁺ and CD34⁺/CXCR4⁺ CPCs were significantly associated with greater WMH volume burden (CD34⁺: β = -0.27, p < 0.01; CD34⁺/CXCR4⁺: β = -0.14, p = 0.03). Higher CD34⁺/VEGFR2⁺ CPCs were associated with higher MoCA scores (β = 0.37, p < 0.01) and language performance on the Boston Naming Test (β = 0.01, p = 0.03) but not with brain phenotypes.

**Conclusions:** Reduced regenerative capacity is associated with worse cognitive performance on global tests and with vascular brain injury, including WMH volume and cortical thinning. If validated in future studies, these findings may highlight regenerative capacity as a promising therapeutic target for mitigating cognitive decline.

## Introduction

Pluripotent stem cells differentiate into lineage-specific progenitor cells that promote tissue repair and regeneration after injury^1^. Bone marrow-derived progenitor cells circulate in peripheral blood, and these rare circulating progenitor cells (CPCs) can be enumerated within the peripheral mononuclear cell population using flow cytometry and various cell surface expression of specific progenitor/stem cell epitopes^2,3^. Clinical studies have generally characterized CPCs using two broad approaches: (i) culture or colony-forming assays, and (ii) flow cytometric identification of mononuclear cells expressing specific surface antigens that enrich for CPC populations^1-3^. CD34 is a prototypical hematopoietic progenitor cell marker, also expressed on endothelial cells, which may explain the vascular tropism and embryonic hemogenic endothelium origin of CD34⁺ cells^2-5^. Co-expression of additional markers, such as CD133 (a transmembrane antigen progenitor marker), CXCR4 (a chemokine receptor involved in homing to injured tissue), and VEGFR2 (a receptor enriched on endothelial progenitor subpopulations), has been used to further delineate subtypes with specific regenerative potential^2,6,7^. In this study, we utilize flow cytometry to quantify CPC-enriched populations of CD34+, CD34+/CD133+, CD34+/CXCR4+, and CD34+/VEGFR2+ cells as measures of endogenous regenerative capacity.

Systemic and cerebral vascular pathologies are recognized as major contributors to cognitive decline and dementia. Vascular brain injury coexists with neurodegenerative pathology, as autopsy studies demonstrate mixed Alzheimer’s and cerebrovascular disease are prevalent in late-life dementia ^8,9^. Among cerebrovascular diseases, cerebral small vessel disease (SVD) is particularly prevalent in older adults and is a key driver of vascular cognitive impairment ^10,11^. Cerebral SVD is recognized as the most prevalent neuroimaging finding in those with vascular cognitive impairment and often manifests as white matter hyperintensities (WMHs), although WMHs are not exclusively vascular in origin^11-13^. Cognitive impairment and cardiovascular disease share overlapping risk factors, and vascular dysfunction is posited as the common pathogenetic injury ^9^. Neuroimaging markers are widely used to risk-stratify and support diagnostic classifications of older adults with cognitive decline. These include WMHs volume, cortical thinning, and global and hippocampal atrophy ^10,12-17^. WMH volume has been associated with poorer executive function and processing speed, while mean whole-brain cortical thickness serves as a sensitive marker of brain aging and gray matter neurodegeneration, with greater thinning observed in individuals experiencing accelerated cognitive decline^18-20^.

In a younger (mean age 49 years), healthier and highly educated population sample, we had previously demonstrated using cognitive scoring tests, that a lower CPC count was associated with a greater decline in cognitive performance over 4 years of longitudinal follow-up, indicating a potentially critical contribution of the endogenous regenerative capacity, reflected by CPC counts to cognitive aging ^21^. We also demonstrated a gradual reduction in CPC counts with aging, a decline that is steeper in those with greater exposure to multiple cardiovascular risk factors or with established cardiovascular disease ^22^. In addition, our previous findings indicate that a worse cardiovascular health profile and abnormal peripheral microvascular function had cognitive decline ^23,24^. Whether CPC counts are associated with neuroimaging markers of cognitive impairment is not fully described. Herein, in an older, clinically at-risk population enriched for vascular risk factors and including individuals with both normal cognition (NC) and consensus diagnosis mild cognitive impairment (MCI), we investigated whether low circulating progenitor cell (CPC) levels are associated with cognitive performance and magnetic resonance imaging (MRI) markers of vascular brain injury. Our overarching hypothesis was that diminished endogenous regenerative potential, reflected by lower CPC counts, is linked to cognitive decline and neurovascular injury patterns in the brain.

## Methods

### Study population

This cross-sectional analysis used data from Brain Stress, Hypertension, and Aging Research Program (B-SHARP), a prospective, single-center program research platform at Emory University designed to investigate vascular and biological contributors to cognitive aging^25^. B-SHARP enrolled community-dwelling adults aged ≥50 years with an emphasis on recruiting racially diverse individuals across the cognitive spectrum. Participants underwent harmonized protocols for cognitive assessments, neuroimaging, blood testing and were subsequently enrolled in observational studies or clinical trials based on their eligibility.

Data were pooled from two harmonized B-SHARP studies: 1) VAScular ContribUtors to prodromaL AlzheimeR’s disease (VASCULAR) — an observational cohort of self-identified Black and White individuals with normal cognition (NC) or mild cognitive impairment (MCI); 2) Cognitive Aging Linked to Biology, Race, and Exposures (CALIBREX; NCT01984164) — a randomized, double-blind trial comparing candesartan versus lisinopril in older adults with hypertension and executive MCI, with a focus on underrepresented minorities to enhance generalizability^23,26^. NC in VASCULAR was defined by a MoCA score ≥26, CDR = 0, and education-adjusted normal performance on the Logical Memory subscale from the Wechsler Memory Scale–Revised^27,28^. MCI classification in VASCULAR required subjective memory complaints, impaired Logical Memory performance, MoCA <26, FAQ <9, and both a global Clinical Dementia Rating (CDR) score of 0.5 and a CDR Memory subscale score of 0.5. CALIBREX defined executive MCI as MoCA ≤26 with impaired performance (≤10th percentile) on ≥1 executive function task. Final diagnoses in both studies were assigned by a multidisciplinary adjudication committee via consensus review of clinical and neuropsychological data. Both studies were conducted at the same location using similar protocols for data collections.

Key exclusion criteria across both studies included prior diagnosis of dementia, clinical stroke within the past 3 years, or comorbid neurological or psychiatric conditions that could confound cognitive testing (e.g., multiple sclerosis, epilepsy, schizophrenia). Demographic data, education (in years), cardiovascular and metabolic health history (e.g., hypertension, diabetes, hyperlipidemia, heart failure, kidney disease) were collected via structured questionnaires and verified through chart abstraction when possible. The Emory University Institutional Review Board approved all study procedures, and all participants provided written informed consent prior to enrollment.

### Cognitive testing

All participants underwent standardized cognitive assessments using validated tools to evaluate multiple domains of cognitive function. Global cognition was assessed using the Montreal Cognitive Assessment (MoCA), a 30-point screening tool that evaluates memory, attention, language, visuospatial skills, and executive function^29^.

Executive function was assessed using the Trail Making Test (TMT), Parts A and B^30^. TMT Part A measures motor speed and visual scanning, while Part B assesses cognitive flexibility and task-switching. To isolate executive function, we calculated the difference score (TMT B − A), which adjusts for motor and perceptual demands. Higher TMT scores indicate longer completion times and suggest impaired executive function^31^. Participants unable to complete TMT Part B within 5 minutes were stopped, and their scores were recorded as 300 seconds.

Verbal and episodic memory were evaluated using the Hopkins Verbal Learning Test–Revised (HVLT-R), which includes delayed recall (20 minutes), retention, and recognition discrimination indices^32^. Language abilities were assessed using the Boston Naming Test (BNT)^33,34^.

MCI diagnoses were adjudicated by study clinicians (IH, FG) following review of cognitive evaluations and clinical history and physical examination. In cases of diagnostic discordance, a third clinician (JL) provided final adjudication through consensus.

### Neuroimaging markers

We examined the relationship between CPC counts and three prespecified MRI-derived markers: total WMH volume, mean of whole-brain cortical thickness, and hippocampal volume (adjusted for intracranial volume). All patients underwent a high-resolution 3T brain MRI using 3D Fast Spoiled Gradient Recalled Echo Sequence. Structural sequences included a 3D Fast Spoiled Gradient-Recalled Echo (FSPGR), 3D T2 Fluid-Attenuated Inversion Recovery (FLAIR) Fast sequences. WMH volume was quantified using a validated morphometric image analysis package (BIANCA - FSL)^13^. Although WMHs are commonly interpreted as markers of cerebral small vessel disease, they may also reflect non-vascular pathology. Greater WMH burden has been associated with accelerated cognitive impairment after adjusting for traditional risk factors and cerebrospinal fluid biomarkers ^12,35^. Hippocampal volumes were derived from high-resolution T1-weighted images and the FREE-SURFER package; left and right hippocampal volumes were summed to obtain total hippocampal volume estimates^36,37^. Intracranial volume was used for normalization to account for interindividual size variability. Cortical thickness, a sensitive marker of brain aging, vascular injury, and neurodegeneration, was calculated from T1-weighted scans using an automated image processing package in FREE_SURFER, averaged across the cerebrum. It reflects the thickness of the cerebral cortex, which governs higher cognitive functions such as memory, attention, perception, and language ^19,20^.

### Flow cytometry and progenitor cell assay

Peripheral venous blood samples were collected in ethylenediaminetetraacetic acid tubes. Flow cytometry was used to quantify CPCs as CD45^med^ mononuclear cells expressing the CD34 epitope ^22,38^. Blood samples were prepared within 4 hours of collection and incubated with fluorochrome-labeled monoclonal antihuman mouse antibodies to identify and enumerate using flow cytometry as CD45med cells co-expressing CD34, CD133, VEGFR2, and CXCR4 epitopes ^38^. Flow cytometry data were analyzed with Flowjo software (Treestar, Ashland, Oregon) with a filter set at CD45^med^ cells. The exclusion of CD45^bright^ cells helped eliminate lymphoblasts, and the exclusion of CD45-cells helped eliminate nonhematopoietic progenitors such as mesenchymal or osteoprogenitor cells. CPC population counts (CD34^+^, CD34^+^/CD133^+^, CD34^+^/CXCR4^+^, and CD34^+^/VEGFR2^+^) were measured using CD45^med+^ filter and reported as counts per µL ^38^. *<u>Reproducibility:</u>* Two technicians analyzed twenty samples on two occasions in our lab. The variability coefficients were 2.9%, 4.8%, 6.5%, and 21.6% for CD34+, CD34^+^/CD133^+^, CD34^+^/CXCR4^+^, CD34^+^/VEGFR2^+^ CPCs, respectively ^22^.

### Statistical Methods

Baseline characteristics of the total sample and sub-groups stratified by cognition (NC vs. MCI) were summarized using proportions for categorical variables and means or medians for continuous variables. Group differences were assessed using chi-squared tests for categorical variables and independent sample *t*-tests for normally distributed continuous variables. CPC counts were non-normally distributed and were log-transformed prior to regression analysis. In addition, WMH volume, mean whole-brain cortical thickness and hippocampal volume, adjusted for intracranial volume, were also log-transformed due to non-normal distribution and to satisfy assumptions of linear regression.

Univariate and multivariable linear regression models were used to examine the cross-sectional associations between log-transformed CPC counts and cognitive performance measures, as well as log-transformed MRI-based markers of vascular brain injury. Sequential models were constructed after including prespecified variables to the unadjusted model (Model 1). In Model 2, we added demographic characteristics (age, sex, race, and years of education). In Model 3, we further adjusted for vascular and metabolic comorbidities, including body mass index and history of hypertension, diabetes mellitus, hyperlipidemia, congestive heart failure, and chronic kidney disease. Beta coefficients and 95% confidence intervals were reported from each model. Logistic regression was additionally used to assess group differences in CPC levels between NC and MCI subgroups. All analyses were performed using R version 4.3.2 (R Foundation for Statistical Computing, Vienna, Austria) ^39^. *P* values of <0.05 from two-sided tests were considered to indicate statistical significance.

For visualization, CPC subtypes were divided into quartiles (Q1–Q4, with Q1 as the reference), and compared against Model 3-adjusted MoCA scores and neuroimaging markers (WMH, cortical thickness, and hippocampal volume). Forest plots summarized Model 3 beta coefficients and p-values for each CPC subtype across VASCULAR, CALIBREX, and pooled cohorts to aid interpretation of direction, precision, and consistency of associations. These plots were constructed using R version 4.3.2 (R Foundation for Statistical Computing, Vienna, Austria)^39^.

## Results

### Demographics and risk factors

A total of 283 participants, 181 from VASCULAR and 102 from CALIBREX, were enrolled. The mean age was 64.7 ± 7.5 years, 59% were women, and 39.2% self-identified as Black **(Table 1, Supplemental Table 1)**. Participants with MCI were significantly older than those with NC, had fewer years of education, higher BMI, and a greater prevalence of hypertension and diabetes mellitus. There were no significant group differences in sex or race, or prevalence of hyperlipidemia, heart failure, or kidney disease. As expected, the NC group, individuals with MCI showed significantly worse cognitive performance across all domains, including lower MoCA scores, slower TMT B–A times, and poorer performance on HVLT-R total recall, delayed recall, retention, and recognition, as well as the Boston Naming Test (*p* < 0.01 for all). Neuroimaging measures revealed that participants with MCI had greater WMH volume (*p* = 0.04), lower mean whole-brain cortical thickness (*p* < 0.01), and smaller hippocampal volumes (*p* < 0.01).

**Table 1:**
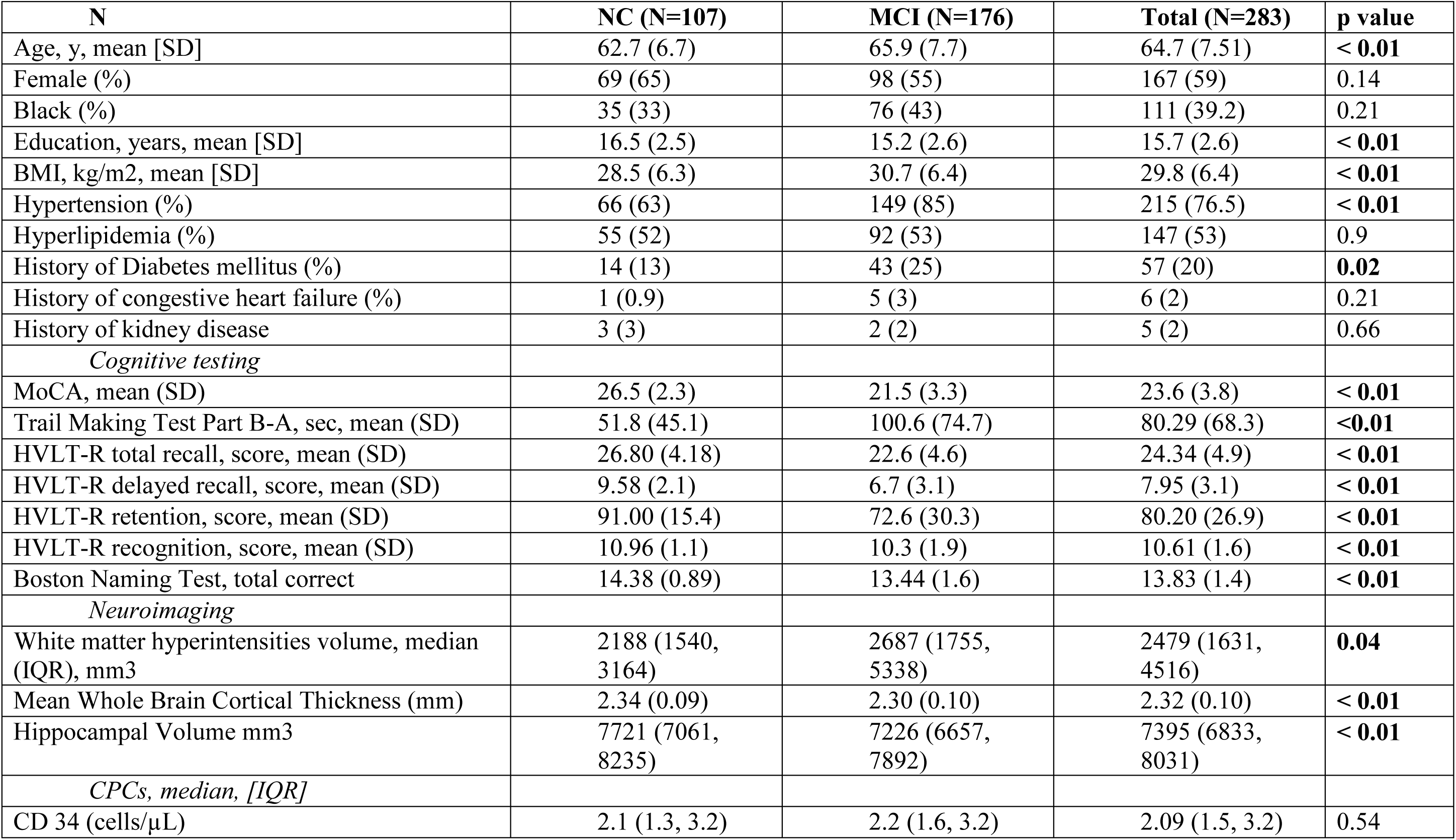
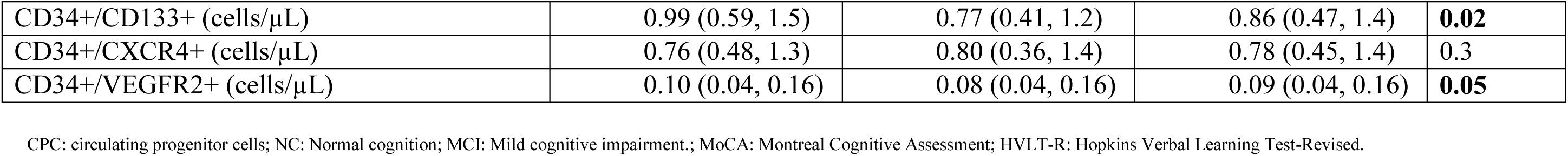
Baseline characteristics of participants with normal cognition (NC) and mild cognitive impairment (MCI) in the pooled cohort.

| **N** | **NC (N=107)** | **MCI (N=176)** | **Total (N=283)** | **p value** |
| --- | --- | --- | --- | --- |
| Age, y, mean [SD] | 62.7 (6.7) | 65.9 (7.7) | 64.7 (7.51) | < **0.01** |
| Female (%) | 69 (65) | 98 (55) | 167 (59) | 0.14 |
| Black (%) | 35 (33) | 76 (43) | 111 (39.2) | 0.21 |
| Education, years, mean [SD] | 16.5 (2.5) | 15.2 (2.6) | 15.7 (2.6) | < **0.01** |
| BMI, kg/m<sup>2</sup>, mean [SD] | 28.5 (6.3) | 30.7 (6.4) | 29.8 (6.4) | < **0.01** |
| Hypertension (%) | 66 (63) | 149 (85) | 215 (76.5) | < **0.01** |
| Hyperlipidemia (%) | 55 (52) | 92 (53) | 147 (53) | 0.9 |
| History of Diabetes mellitus (%) | 14 (13) | 43 (25) | 57 (20) | **0.02** |
| History of congestive heart failure (%) | 1 (0.9) | 5 (3) | 6 (2) | 0.21 |
| History of kidney disease | 3 (3) | 2 (2) | 5 (2) | 0.66 |
| *Cognitive testing* |  |  |  |  |
| MoCA, mean (SD) | 26.5 (2.3) | 21.5 (3.3) | 23.6 (3.8) | < **0.01** |
| Trail Making Test Part B-A, sec, mean (SD) | 51.8 (45.1) | 100.6 (74.7) | 80.29 (68.3) | < **0.01** |
| HVLT-R total recall, score, mean (SD) | 26.80 (4.18) | 22.6 (4.6) | 24.34 (4.9) | < **0.01** |
| HVLT-R delayed recall, score, mean (SD) | 9.58 (2.1) | 6.7 (3.1) | 7.95 (3.1) | < **0.01** |
| HVLT-R retention, score, mean (SD) | 91.00 (15.4) | 72.6 (30.3) | 80.20 (26.9) | < **0.01** |
| HVLT-R recognition, score, mean (SD) | 10.96 (1.1) | 10.3 (1.9) | 10.61 (1.6) | < **0.01** |
| Boston Naming Test, total correct | 14.38 (0.89) | 13.44 (1.6) | 13.83 (1.4) | < **0.01** |
| *Neuroimaging* |  |  |  |  |
| White matter hyperintensities volume, median (IQR), mm<sup>3</sup> | 2188 (1540, 3164) | 2687 (1755, 5338) | 2479 (1631, 4516) | **0.04** |
| Mean Whole Brain Cortical Thickness (mm) | 2.34 (0.09) | 2.30 (0.10) | 2.32 (0.10) | < **0.01** |
| Hippocampal Volume mm<sup>3</sup> | 7721 (7061, 8235) | 7226 (6657, 7892) | 7395 (6833, 8031) | < **0.01** |
| *CPCs, median, [IQR]* |  |  |  |  |
| CD 34 (cells/μL) | 2.1 (1.3, 3.2) | 2.2 (1.6, 3.2) | 2.09 (1.5, 3.2) | 0.54 |
| CD34+/CD133+ (cells/μL) | 0.99 (0.59, 1.5) | 0.77 (0.41, 1.2) | 0.86 (0.47, 1.4) | **0.02** |
| CD34+/CXCR4+ (cells/μL) | 0.76 (0.48, 1.3) | 0.80 (0.36, 1.4) | 0.78 (0.45, 1.4) | 0.3 |
| CD34+/VEGFR2+ (cells/μL) | 0.10 (0.04, 0.16) | 0.08 (0.04, 0.16) | 0.09 (0.04, 0.16) | **0.05** |

CPC: circulating progenitor cells; NC: Normal cognition; MCI: Mild cognitive impairment.; MoCA: Montreal Cognitive Assessment; HVLTL-R: Hopkins Verbal Learning Test-Revised.

### Relationship between CPCs and Cognitive scores

In fully adjusted models (Model 3), lower CD34⁺/CD133⁺ and CD34⁺/VEGFR2⁺ cell counts were independently associated with lower MoCA scores (β = 0.59, *p* = 0.01 and β = 0.37, *p* < 0.01, respectively) **(Table 2a).**

**Table 2:** Relationship between CPC counts and (a) MoCA, (b) WMH, (c) Mean Whole Brain Cortical thickness, (d) Hippocampal Volume (adjusted for intracranial volume).

| CPC | Model 1 |  | Model 2 |  | Model 3 |  |
| --- | --- | --- | --- | --- | --- | --- |
| | $\beta$ | p-value | $\beta$ | p-value | $\beta$ | p-value |
| CD34<sup>+</sup> | -0.49 | 0.20 | 0.36 | 0.30 | -0.19 | 0.60 |
| CD34<sup>+</sup>/CD133<sup>+</sup> | 0.45 | 0.08 | 0.50 | **0.03** | 0.59 | **0.01** |
| CD34<sup>+</sup>/CXCR4<sup>+</sup> | 0.24 | 0.34 | 0.23 | 0.33 | 0.23 | 0.32 |
| CD34<sup>+</sup>/VEGFR2<sup>+</sup> | 0.41 | **< 0.01** | 0.40 | **< 0.01** | 0.37 | **< 0.01** |

**(b) (Log) White Matter Hyperintensity Volumes:**
| CPC | Model 1 |  | Model 2 |  | Model 3 |  |
| --- | --- | --- | --- | --- | --- | --- |
| | $\beta$ | p-value | $\beta$ | p-value | $\beta$ | p-value |
| CD34<sup>+</sup> | -0.05 | 0.49 | -0.08 | 0.28 | -0.27 | **< 0.01** |
| CD34<sup>+</sup>/CD133<sup>+</sup> | -0.05 | 0.32 | -0.03 | 0.44 | -0.15 | **0.01** |
| CD34<sup>+</sup>/CXCR4<sup>+</sup> | -0.13 | **0.01** | -0.09 | 0.06 | -0.14 | **0.03** |
| CD34<sup>+</sup>/VEGFR2<sup>+</sup> | -0.03 | 0.19 | -0.01 | 0.53 | -0.05 | 0.19 |

**(c) Mean Whole-Brain Cortical Thickness:**
| CPC | Model 1 |  | Model 2 |  | Model 3 |  |
| --- | --- | --- | --- | --- | --- | --- |
| | $\beta$ | p-value | $\beta$ | p-value | $\beta$ | p-value |
| CD34<sup>+</sup> | 0.01 | 0.61 | 0.01 | 0.38 | 0.01 | 0.19 |
| CD34<sup>+</sup>/CD133<sup>+</sup> | 0.01 | 0.27 | 0.01 | 0.31 | 0.01 | **0.01** |
| CD34<sup>+</sup>/CXCR4<sup>+</sup> | 0.01 | 0.15 | 0.01 | 0.38 | 0.01 | 0.29 |
| CD34<sup>+</sup>/VEGFR2<sup>+</sup> | 0.01 | 0.30 | **< 0.01** | 0.36 | **< 0.01** | 0.12 |

**(d) (Log) Mean Hippocampal Volume (adjusted for intracranial volume):**
| **CPC** | **Model 1** |  | **Model 2** |  | **Model 3** |  |
| --- | --- | --- | --- | --- | --- | --- |
|  | **$\beta$** | **p-value** | **$\beta$** | **p-value** | **$\beta$** | **p-value** |
| CD34<sup>+</sup> | 0.01 | 0.46 | 0.01 | 0.29 | 0.01 | 0.25 |
| CD34<sup>+</sup>/CD133<sup>+</sup> | 0.01 | 0.17 | 0.01 | 0.14 | 0.01 | 0.21 |
| CD34<sup>+</sup>/CXCR4<sup>+</sup> | 0.01 | 0.39 | 0.01 | 0.57 | 0.01 | 0.08 |
| CD34<sup>+</sup>/VEGFR2<sup>+</sup> | -0.01 | 0.39 | -0.01 | 0.78 | -0.01 | 0.71 |

Model 1: Univariate. Model 2 was adjusted for continuous age, sex, race, and education level (in years). Model 3 adjusted for Model 2 + body mass index, hypertension, diabetes mellitus, hyperlipidemia, congestive heart failure, and chronic kidney disease.

For other cognitive domains assessed only higher CD34⁺/VEGFR2⁺ counts were associated with better performance on the Boston Naming Test (β = 0.01, *p* = 0.03) **(Supplemental Table 2a-f)**.

### Relationship between CPCs and Neuroimaging markers

In fully adjusted linear regression models (Model 3), lower levels of CD34⁺, CD34⁺/CD133⁺, and CD34⁺/CXCR4⁺ cells were significantly associated with greater white matter hyperintensity (WMH) volumes (p = 0.01, 0.01, and 0.03, respectively), indicating a link between reduced regenerative capacity and cerebrovascular injury **(Table 2b)**. Lower levels of CD34⁺/CD133⁺ cells were also associated with reduced mean whole-brain cortical thickness after full adjustment (p = 0.01), suggesting the potential role of reduced regenerative capacity in cortical atrophy **(Table 2c**, **Figure 1)**. CPC counts were not associated with hippocampal volume adjusted for intracranial volume **(Table 2d)**. When divided into quartiles, individuals in the highest CD34⁺/CD133⁺ CPC quartile (Q4) exhibited approximately a 2-point higher adjusted MoCA score, 36% lower adjusted WMH volumes, and 0.06 mm greater mean whole-brain cortical thickness compared to those in the lowest quartile (Q1) **(Figure 1).**

**Figure 1:**
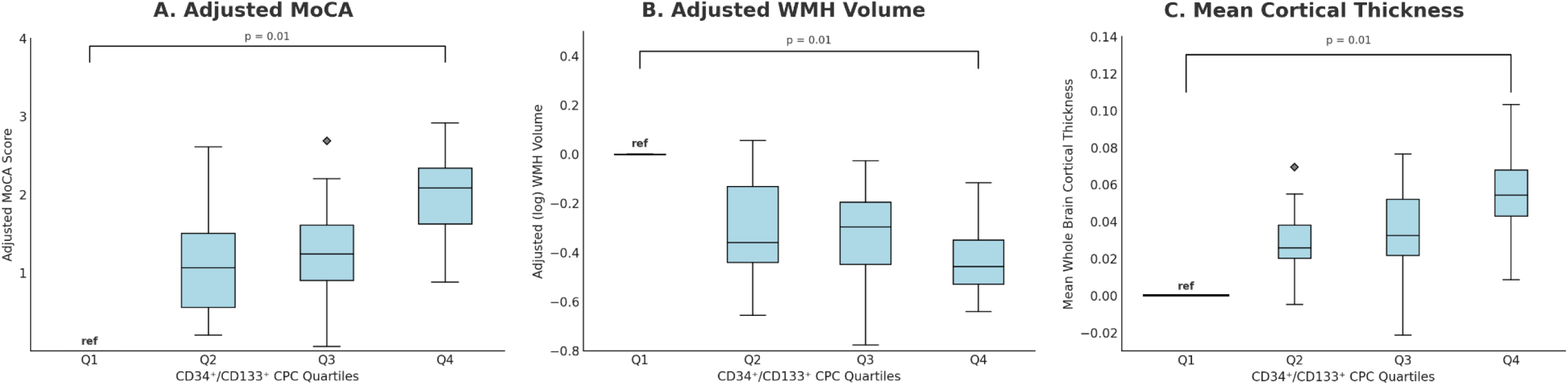
Associations between CD34⁺/CD133⁺ CPC quartiles and cognitive and neuroimaging measures. Boxplots display fully adjusted (Model 3) values across quartiles (Q1–Q4) of log-transformed CD34⁺/CD133⁺ CPC levels, with Q1 (lowest) as the reference. (A) Higher CD34⁺/CD133⁺ CPC quartiles were associated with higher adjusted MoCA scores (p = 0.01). (B) Higher quartiles were associated with lower log-transformed WMH volumes (p = 0.01). (C) Higher quartiles were associated with greater mean whole-brain cortical thickness (p = 0.01). Models adjusted for age, sex, race, education, BMI, hypertension, diabetes, hyperlipidemia, congestive heart failure, and chronic kidney disease.

Findings were directionally similar in the VASCULAR and CALIBREX cohorts, although associations reached statistical significance primarily within the VASCULAR cohort, likely due to its larger sample size **(Supplemental Figure 1).**

### Relationship between CPCs and Mild Cognitive Impairment (MCI)

In univariate logistic regression, lower levels of CD34⁺/CD133⁺ (OR = 0.72, 95% CI: 0.53–0.96, p = 0.02) and CD34⁺/VEGFR2⁺ (OR = 0.55, 95% CI: 0.42–0.71, p < 0.01) were associated with higher odds of MCI, though these associations attenuated and became non-significant after multivariate adjustment **(Supplemental Table 3)**. Lower CD34⁺/CXCR4⁺ levels showed a non-significant trend toward higher MCI odds (unadjusted OR = 0.78, p = 0.08; adjusted OR = 0.76, p = 0.23).

## Discussion

Novel findings of our study are that lower levels of CPCs, reflective of reduced vascular regenerative capacity, are associated with lower cognitive performance on the global measure, lower mean whole brain cortical thickness and greater burden of WMH volumes. We employed a comprehensive battery of cognitive tests and MRI measures to rigorously study the relationship between CPC levels and markers of cognitive impairment, including WMH volume and cortical thinning. Goldstein et al. have demonstrated that a greater WMH burden and cortical thinning, but not the hippocampal volumes, were related to subjective cognitive complaints in older adults suggesting that vascular brain injury is likely an important contributor to cognitive manifestation of brain illnesses ^40^. Our previous study in a community-based cohort of healthy adults also indicated that higher CPC counts were associated with a lower risk of cognitive decline in multiple cognitive domains during longitudinal follow-up ^21^. In another cross-sectional analysis of patients with coronary artery disease, we showed that individuals with lower levels of CPCs had worse memory performance and cognitive function ^41^. Our current findings are also supported by a previous study by Nation et al. that lower CD34⁺ levels correlated with worse memory performance and reduced mean whole brain cortical thickness ^42^. Notably, they did not find a relationship between CPCs and WMH volume, likely due to a smaller sample size and because their participants were healthier with minimal small-vessel disease burden ^42^. Although prior studies have reported inconsistent associations, our larger and more diverse cohort demonstrated a robust relationship between lower CPC counts and both cognition and brain WMH volume burden, indicating that CPC depletion is linked to greater cerebral small vessel disease. Furthermore, in contrast to prior studies, our study was able to confirm that regenerative capacity is related to brain outcomes independently of other CV risks and comorbidities.

Notably, in our study, CD34⁺/CD133⁺ CPCs demonstrated the most consistent and robust associations across structural and cognitive measures, suggesting this subset may be particularly relevant in brain aging **(Central Illustration)**. CD34⁺/CD133⁺ CPCs represent an early-stage hematopoietic progenitor cell population, reflecting a pool of cells with high proliferative and regenerative potential that diminishes with aging and vascular risk exposure^2,6^. This subtype’s strong correlation with cortical thinning, WMH burden, and MoCA scores in our study underscores its potential as a sensitive biomarker of impaired neurovascular repair capacity.

In our study, lower CPC levels were associated with higher odds of MCI in unadjusted models, but these associations attenuated and became non-significant after adjustment for demographic and vascular risk factors, suggesting that while CPC depletion may contribute to cognitive decline, CPC levels are more closely linked to subclinical neuroimaging markers and continuous cognitive measures than to categorical MCI diagnosis, which can be influenced by multiple non-vascular factors. Supporting this interpretation, a prior study using a colony-forming assay found fewer hematopoietic progenitor cell colonies in individuals with MCI, along with worse memory, executive, and language performance, further strengthening the concept that impaired regenerative capacity is linked to, and may contribute to, cognitive decline^43^.

Age-related declines in CPC counts can be significantly accentuated by cardiovascular risk factors ^22^, which also appears to contribute to cognitive decline ^44^. Lower CPC counts, representative of impaired regenerative capacity, may predispose to rapidly progressive microvascular disease, endothelial dysfunction, reduced repair and angiogenesis, increased blood-brain barrier permeability, and ultimately greater ischemic injury ^45-47^. Over time, such processes manifest as extensive WMH and brain atrophy, which in turn relate to cognitive impairment. WMH volumes have also been linked with chronic inflammation, and we have previously shown that a lower CPC count portends a reduced ability to mitigate inflammation, leading to white matter damage and cognitive impairment ^13,48^. Since older adults often exhibit a mixed dementia phenotype, replenishing progenitor cells may also help limit neurodegeneration by improving cerebral perfusion, increasing endothelial amyloid clearance and improving neurovascular coupling ^49,50^.

Experimental studies have previously demonstrated the crucial contribution of CPCs to neurogenesis and cognitive function ^41,42^. Kapoor et al showed that higher CPC counts attenuate WMH burden in APOE4 carriers, suggesting a protective role of CPCs on cerebral microvasculature. Progenitor cells enhance neurogenesis and angiogenesis in mice after an ischemic stroke ^51^. Transgenic mice undergoing hippocampal injection of CPCs had improvement in learning and memory function and upregulation in blood-brain barrier tight junction proteins, increases in micro-vessel density, and decreased plaque deposition and neuronal apoptosis ^52^. Mobilization of progenitor cells using granulocyte monocyte colony-stimulating factor improved memory and synapse formation ^53^. Clinical trials in individuals with Alzheimer’s dementia using GM-CSF analogs are currently underway ^54^. Collectively, these studies highlight the therapeutic potential of progenitor cells in improving vascular health and cognitive function in models of Alzheimer’s disease.

### Strengths and Limitations

Key strengths of our study is its size after combining two complementary cohorts with identical phenotyping that enhanced population diversity and statistical power. The findings were directionally similar within the two cohorts and strengthen the generalizability and internal validity of our findings. We employed standardized CPC quantification with flow cytometry and rigorous cognitive and MRI phenotyping. We examined multiple biologically relevant CPC subsets enriched for hematopoietic and endothelial progenitor activity, and our modeling approach accounted for a range of demographic and cardiovascular risk factors. However, the cross-sectional nature of our analysis precludes causal inference. Future research should investigate the long-term consequences of low CPC counts on the progression of cognitive impairment. Additionally, our findings highlight the need for further studies to explore specific progenitor subpopulations and identify cell types and regulatory factors relevant to cognitive impairment.

## Conclusions

Impaired endogenous vascular regenerative capacity, assessed as lower levels of CPCs, was independently associated with structural and functional markers of cognitive impairment and vascular brain injury. Longitudinal studies examining the decline in cognition in relation to low CPC counts and studies using progenitor or stem cell therapy for cognitive decline are warranted.

## Data Availability

The data that support the findings of this study are not publicly available due to privacy and institutional restrictions. Deidentified participant data may be available from the corresponding author upon reasonable request and with appropriate institutional approvals.

